# Quality of maternal and newborn care, perinatal mental health and the emotional birth experience of women: findings of the IMAgiNE EURO study in Belgium

**DOI:** 10.64898/2025.12.19.25342646

**Authors:** Anna Galle, Julie Verschueren, Victoria Vercaempst, Maria Verdecchia, Ilaria Mariani, Lorenzo Giovanni Cora, Arianna Bomben, Margherita Camanni, Mieke Embo, Nele Vaerewijck, Marza Lazzerini, the IMAgiNE EURO Study Group

## Abstract

**Background:** In Belgium, maternal and newborn health indicators indicate high quality of care compared to other countries of Europe. However, some challenges still persist in the quality of maternal and newborn care (QMNC) and little is known regarding women’s experience with care and their mental health (MH).

**Methods:** We conducted a cross-sectional survey among 621 women who gave birth between March 2022 and January 2025 in Belgium, using the WHO-based IMAgiNE EURO questionnaire, updated with MH items. Descriptive statistics summarized QMNC and MH outcomes, while multivariate logistic regression examined factors associated with negative emotional childbirth experiences.

**Results:** The median QMNC index was 260/300, suggesting high adherence to WHO quality standards. However, 33.5% of respondents reported ineffective communication, 34.0% a lack of involvement in decision-making, 8.7% no emotional support from health care providers, and 8.2% reported abuse. Overall, 40.9% of women reported emotional difficulties related to childbirth, with 10% reporting a negative impact on wellbeing. MH screening and support were not structurally embedded in perinatal care. Emergency caesarean sections (aOR 14.94, 95%CI), instrumental births (aOR 2.36), fundal pressure (aOR 4.11), and abuse (aOR 3.74) were significantly associated with emotional difficulties around childbirth. Protective factors included higher QMNC scores (aOR 0.97) and the presence of an obstetric consultant (aOR: 0.44).

**Conclusion:** Belgium shows a high level of QMNC, yet significant gaps remain in communication, MH screening and support, and adherence to evidence-based practices. Our study highlights a strong association between certain childbirth interventions and a negative emotional birth experience, emphasizing the need for further research into the underlying causal patterns and contextual factors shaping women’s childbirth experiences.

**Strengths and Limitations:**

- The study used a validated WHO standards–based questionnaire, ensuring comparable measurement of quality of care indicators.
- The updated questionnaire incorporated measures of emotional childbirth experience and perinatal mental health, addressing an important evidence gap in Belgian maternity care research.
- The overrepresentation of Dutch-speaking and highly educated women may limit applicability to the entire Belgian population.
- Participation was voluntary and data were self-reported, potentially leading to over- or underestimation of key outcomes, especially for sensitive topics such as mental health difficulties and experiences of abuse.

## BACKGROUND

The perinatal period is a unique phase in people’s lives, encompassing pregnancy, childbirth, and the immediate postpartum period, and is marked by significant biological and psychological changes. High-quality maternal and newborn care is essential in this time and considered the cornerstone of a well-functioning healthcare system [1]. Belgium has a robust healthcare system with overall good maternal and newborn health outcomes (including low maternal morbidity and mortality), but several challenges persist. Regional registry data show high variations in clinical practices during childbirth (e.g. caesarean sections, inductions and episiotomies) across hospitals and regions, underscoring the need for more standardized, evidence-based approaches [2–5].

According to the WHO Framework for Quality of Care in maternal and newborn health, both the provision of care and the experience of care are considered as fundamental components of high-quality care [6]. However, data on women’s experience with maternal and newborn health care are rather scarce in Belgium compared to neighbouring countries such as France, the Netherlands and Germany [6, 7]. Also, aspects of mental health in the perinatal period and the emotional birth experience remain underexamined, representing a significant blind spot in research on quality of maternal and newborn health care. Mental health is a critical aspect of women’s well-being, especially during the perinatal period. Research shows that women are particularly vulnerable to MH challenges during this time, with conditions such as perinatal depression, anxiety, and post-traumatic stress disorder (PTSD) affecting around one in five new mothers [8–10]. Notably, quality of care during childbirth plays an important role in the emotional childbirth experience and MH of women. Several studies showed how a negative childbirth experience contributes to maternal depression and anxiety [11–13]. Furthermore, it is estimated that one in three births is perceived as psychologically traumatic, with approximately 4% of women and 1% of their partners developing PTSD as a result [8–10]. MH conditions can have profound effects not only on the mothers’ health but also on the infant and the broader family dynamic. Untreated perinatal MH issues, for example, are associated with adverse outcomes in children, including poor maternal-infant bonding, developmental delays, and increased risks of long-term emotional and behavioural problems [14–17].

Both WHO guidelines and national guidelines (such as the NICE guideline in the UK and KCE guideline in Belgium) recommend MH screening for all mothers during pregnancy and in the postpartum period [18, 19]. Moreover, the WHO advocates for an integration of psychosocial interventions in early childhood health and development programs to improve maternal and child health [20]. In Belgium, a perinatal MH care pathway has been established, including a screening tool and implementation guideline [21, 22]. According to the guideline, every woman should be screened for depression and anxiety by a healthcare provider (HCP) in the perinatal period, preferably around 20 weeks of gestational age and repeated at 6 weeks post-partum. However, as in other European countries, the effective implementation and evaluation of this guideline face several challenges, including fragmented perinatal care, a shortage of trained HCPs, and inadequate funding [23].

IMAgiNE EURO is a multi-country project that began at the onset of the COVID-19 pandemic and is still ongoing today. Through two online surveys, it investigates the perspectives of women and HCPs on the quality of maternal and newborn care (QMNC) around childbirth in hospital settings. The first round of IMAgiNE EURO data collection among women in Belgium (2022-2022) highlighted several gaps, including inadequate and/or unclear communication from HCPs, lack of involvement in choices, inadequate staff number, frequent performance of episiotomies and inadequate pain management [24]. The second round of data collection in Belgium was conducted in the post COVID-19 period (2024-2025), using the same questionnaire on QMNC, but with an updated section on emotional birth experience and perinatal MH.

This study explores the QMNC in Belgium in the post COVID-19 period, with a particular focus on emotional birth experience and perinatal MH. Our first aim was to explore the quality of care around childbirth and current gaps in perinatal MH care provision, and as a secondary objective, we examined the association between women’s sociodemographic and birth characteristics, QMNC and the emotional birth experience.

## METHODS

### Study design and participants

This is a cross-sectional study and is reported according to the Strengthening the Reporting of Observational Studies in Epidemiology (STROBE) guidelines, see additional file 1 [25]. This study included women aged 18 years and older who gave birth between March 2022 and January 2025 in a birth facility in Belgium.

### Data collection

We used a structured online questionnaire to collect data, recorded with Research Electronic Data Capture (REDCap 8.5.21) (Harris et al., 2009) via a centralized platform. The process of questionnaire development, validation, and prior application has been reported elsewhere [26]. The questionnaire included 30 questions, each focusing on a WHO quality measure, spread evenly over three domains of the WHO standards: provision of care, experience of care and availability of human and physical resources [26]. The 30 WHO quality measures contributed to a composite QMNC index, ranging from 0 to 100 for each of the three domains (provision of care, experience of care, resources), for an overall QMNC index ranging from 0 to 300 points, and higher scores indicating higher adherence to WHO Standards [26]. The QMNC index measured quality of hospital-based care, until discharge from the hospital, and was therefore not suitable for women giving birth in other settings.

The second part of the questionnaire was developed in 2024. Five new questions on emotional birth experience and MH– including one open-ended question – were added, exploring key aspects of MH in the perinatal period, as recommended by WHO [26]. The added questions can be found as an additional file (see additional file 2-English Version of the IMAgiNE EURO questionnaire). Questions on participants’ individual characteristics (e.g., socioeconomic background, parity) remained unchanged across both rounds of data collection. Adaptation of the original questionnaire for the second round of data collection was minor and therefore did not include a validation study. The online questionnaire was disseminated in Belgium by social media (Facebook and X) and by distributing leaflets in midwifery practices, maternity wards, postnatal clinics and creches. An anonymized overview of the dissemination sites can be found as an additional file (see additional file 3). Dissemination materials were available in Dutch and English. The Dutch leaflet directed participants immediately to the online survey through a link and a QR code, while the English flyer opened a landing page where participants could choose from 21 languages, including French and German. Belgium consists of three regions: Flanders (Dutch-speaking), Wallonia (French-speaking, with a small German-speaking area), and the Brussels-Capital Region (bilingual French-Dutch). Dissemination efforts were concentrated in the Flanders region, driven by the accessibility of local networks, institutions, and community organizations that supported the distribution of the survey.

### Data analysis

A minimum required sample size of 498 women was calculated, based on preliminary data from previous IMAgiNE EURO publications, on the hypothesis of an average QMNC index of 75% +/-5% (225 +/-15 points, out of 300) and confidence level of 99%. The same sample was adequate on the hypothesis of a proportion of women experiencing difficulties during birth of 33%+/-5% and confidence level of 95% [27]. Cases with >20% missing values were removed, and suspected duplicates - identified through date and place of birth, as well as socio-demographic and obstetric data - were reviewed, and the most recent record was retained.

We calculated absolute frequencies and percentages for sociodemographic variables, the 30 WHO quality measures, and variables related to MH. Quality measures were also presented by labour experience by differentiating between women who experienced labour and those who did not experience labour. The QMNC index was presented as the median and interquartile range (IQR) due to its non-normal distribution both for the overall sample and stratified by domain of care. Only women who completed all 30 quality measures were included in the calculation of the overall QMNC index, while the QMNC index by domain of care was calculated for women who answered all 10 key quality measures of the domain of care under analysis.

In addition, we performed a multivariate logistic regression model with emotional difficulties at birth as the dependent variable. Women who reported their birth experience as negative– whether or not it affected their MH – were combined into a single category. Maternal age, parity, education, migration background, mode of birth, experience of abuse during childbirth (dichotomized for no abuse and any experience of abuse, the latter including “sometimes” or “always” responses in the same category), interventions during childbirth (episiotomy and fundal pressure), the overall QMNC index, and presence of an obstetric consultant or obstetric doctor in post-graduate training directly assisting childbirth as independent variables. Women with missing values for the variables included in the model as well as women with a stillbirth, women admitted to intensive care or their baby admitted to a neonatal intensive care unit were excluded from the model; since their emotional birth experience might be highly affected by these events. For the variable education, the “none”, “primary education” categories were merged with the “GCSE (or equivalent)” category due to their low sample numerosity (<5 women per category). The reference categories for the variables included in the model were selected based on those reporting the highest number of observations, with the exception of the presence of an obstetric consultant at birth, where the absence of an obstetric consultant at birth was selected as reference category for ease of interpretation. A list of the reference categories for the ordinal logistic regression model can be found in additional file 4-Table 1. 95% Wald confidence intervals (CI) were calculated for the odds ratios. Statistical analyses were performed using Stata version 14 (Stata Corporation) and R version 4.1.1 (R Core Team, 2013).

**Table 1.** Characteristics of the respondents (N=621)

| **Gave birth in the same country where they were born** | **N** | **%** |
| --- | --- | --- |
| Yes | 565 | 90.98 |
| No | 50 | 8.05 |
| Missing | 6 | 0.97 |
| **Region** |  |  |
| Brussels | 35 | 5.64 |
| Flanders | 575 | 92.59 |
| Wallonia | 0 | 0 |
| Missing | 11 | 1.77 |
| **Age (years)** |  |  |
| 18-24 | 18 | 2.90 |
| 25-30 | 205 | 33.01 |
| 31-35 | 303 | 48.79 |
| 36-39 | 73 | 11.76 |
| ≥40 | 17 | 2.74 |
| Missing | 5 | 0.81 |
| **Education** |  |  |
| None | 2 | 0.32 |
| Primary education | 4 | 0.64 |
| GCSE (or equivalent) | 101 | 16.26 |
| Vocational qualification or A level (or equivalent) | 231 | 37.20 |
| First degree | 116 | 18.68 |
| Higher degree (Post-graduate degree/master/doctorate or higher) | 162 | 26.09 |
| Missing | 5 | 0.81 |
| **Birth mode** |  |  |
| Non-instrumental vaginal birth | 436 | 70.21 |
| Instrumental vaginal birth | 82 | 13.20 |
| Emergency caesarean section during labour | 40 | 6.44 |
| Emergency caesarean section before going into labour | 22 | 3.54 |
| Planned or elective caesarean section before going into labour | 41 | 6.60 |
| **Type of hospital** |  |  |
| Private hospital | 50 | 8.05 |
| Public hospital | 557 | 89.69 |
| Missing | 14 | 2.25 |
| **Type of healthcare providers who directly assisted birth** |  |  |
| Midwife | 578 | 93.08 |
| Nurse | 141 | 22.71 |
| Student (before graduation) | 224 | 36.07 |
| Obstetrics doctor in post-graduate training (registrar, SHO) | 223 | 35.91 |
| Obstetric consultant | 522 | 84.06 |
| I don't know (healthcare provider did not introduce themselves) | 25 | 4.03 |
| Other | 51 | 8.21 |
| **Language in which questionnaire was filled** |  |  |
| Dutch | 610 | 98.23 |
| Greek | 1 | 0.16 |
| Italian | 1 | 0.16 |
| Latvian | 2 | 0.32 |
| Lithuanian | 1 | 0.16 |
| Norwegian | 3 | 0.48 |
| Polish | 1 | 0.16 |
| Portuguese | 1 | 0.16 |
| Slovenian | 1 | 0.16 |
| Swedish | 1 | 0.16 |
| **Newborn admission to NICU** |  |  |
| Yes | 52 | 8.37 |
| No | 569 | 91.63 |
| **Maternal admission to ICU** |  |  |
| Yes | 12 | 1.93 |
| No | 598 | 96.30 |
| Missing | 11 | 1.77 |
| **Multiple births** |  |  |
| Yes | 12 | 1.93 |
| No | 598 | 96.30 |
| Missing | 11 | 1.77 |
| **Stillbirth** |  |  |
| Yes | 4 | 0.64 |
| No | 606 | 97.58 |
| Missing | 11 | 1.77 |
| **Parity** |  |  |
| 1 | 397 | 63.93 |
| ≥2 | 224 | 36.07 |

Abbreviations: GCSE=General Certificate of Secondary Education. SHO=Senior House Officer. NICU=Neonatal Intensive Care Unit.
ICU=Intensive Care Unit

### Patient and Public Involvement

Patients or the public were not involved in the design, conduct, reporting, or dissemination plans of our research.

### Ethical aspects

The international study was approved by the Institutional Review Board of the coordinating center: the IRCCS “Burlo Garofolo” Trieste (IRB-BURLO 05/2020 15.07.2020) and the Commision Medical Ethics UZ Ghent (THE-2023-0075). The study was conducted according to General Data Protection Regulation requirements and participation in the online survey was voluntary and anonymous. Prior to participation, women were informed of the objectives and methods of the study, including their rights in declining participation, and each participant provided informed consent before responding to the questionnaire. Anonymity in data collection during the survey phase was ensured by not collecting any information that could disclose participant identity, such as facility of birth or day of birth of the woman, while data transmission and storage were secured by encryption.

## RESULTS

### Sociodemographic characteristics

A total of 621 women who gave birth in Belgium between March 2022 and January 2025 participated in the study (see Figure 1). The majority of respondents gave birth in the Flanders region (92.6%) and in the same country where they were born (91.0%) (see table 1). Over 80% of women completed at least a vocational qualification or higher. A large majority (81.80%) of women was between 25-35 with 33.0% of women being between 25-30 years old and 48.79% of women being between 31-35 years old. The most common mode of birth was non-instrumental vaginal birth (70.2%), followed by caesarean section (16.6%) and instrumental vaginal birth (13.2%). In total, 8.4% of women reported their newborn was admitted to the neonatal intensive care unit and 37% of women were multiparous. Most women filled in the questionnaire in the Dutch language (98.23%).

**Figure 1.**
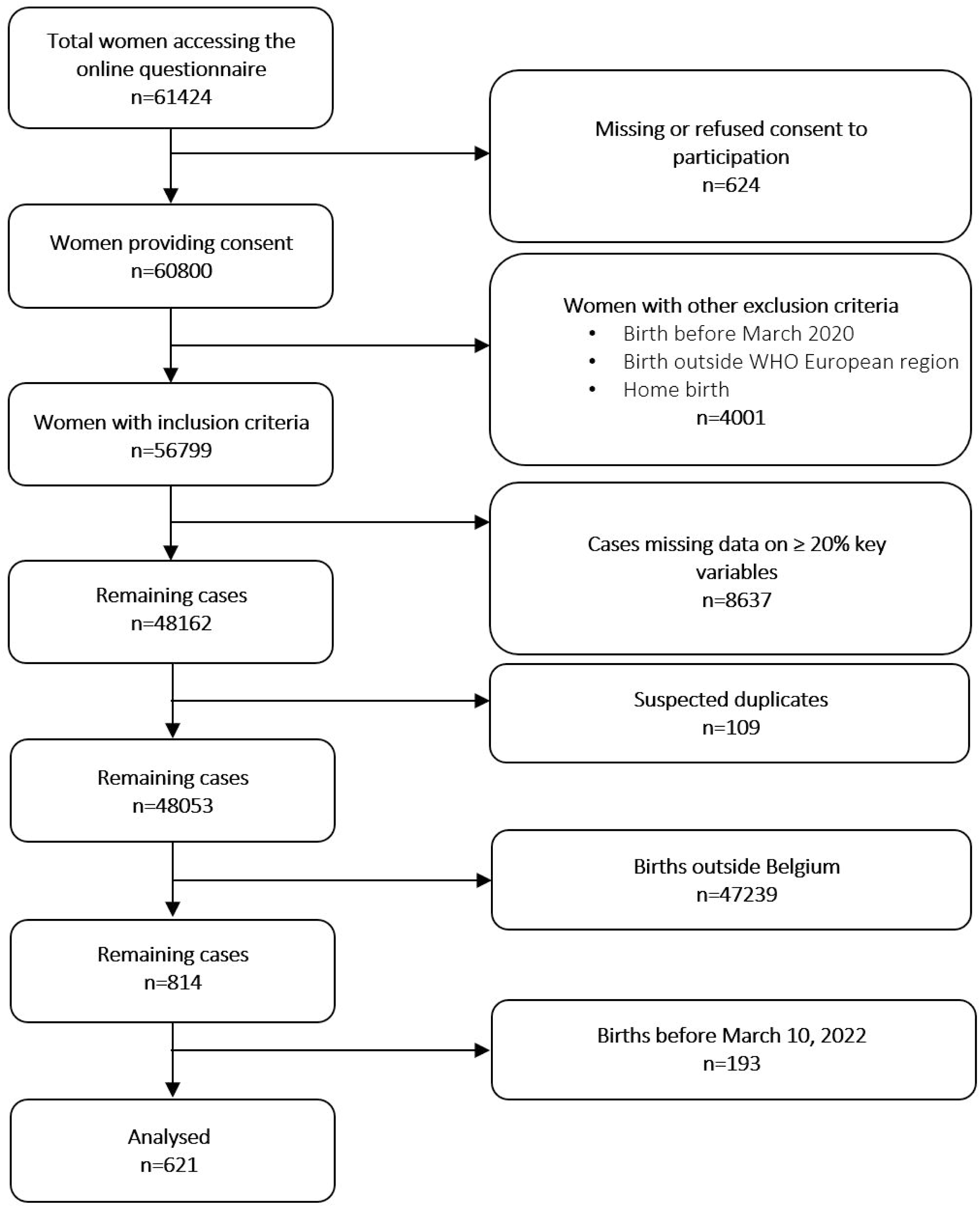
Flow chart of the final study sample.

### Quality of Care

The median overall QMNC index was 260 (IQR 235-275) out of a maximum score of 300, indicating a generally high level of adherence to WHO quality standards (see additional file 4-Table 2). When stratified by the three domains of care, the median index was 90 out of a maximum score of 100 for both provision of care and experience of care (IQRs: 80-95 and 75-100, respectively), and 80 for availability of human and physical resources (IQR: 70-90). Frequencies of all WHO quality measures participating in the QMNC index can be found in Figure 2 and additional file 4-Table 3.

**Figure 2.**
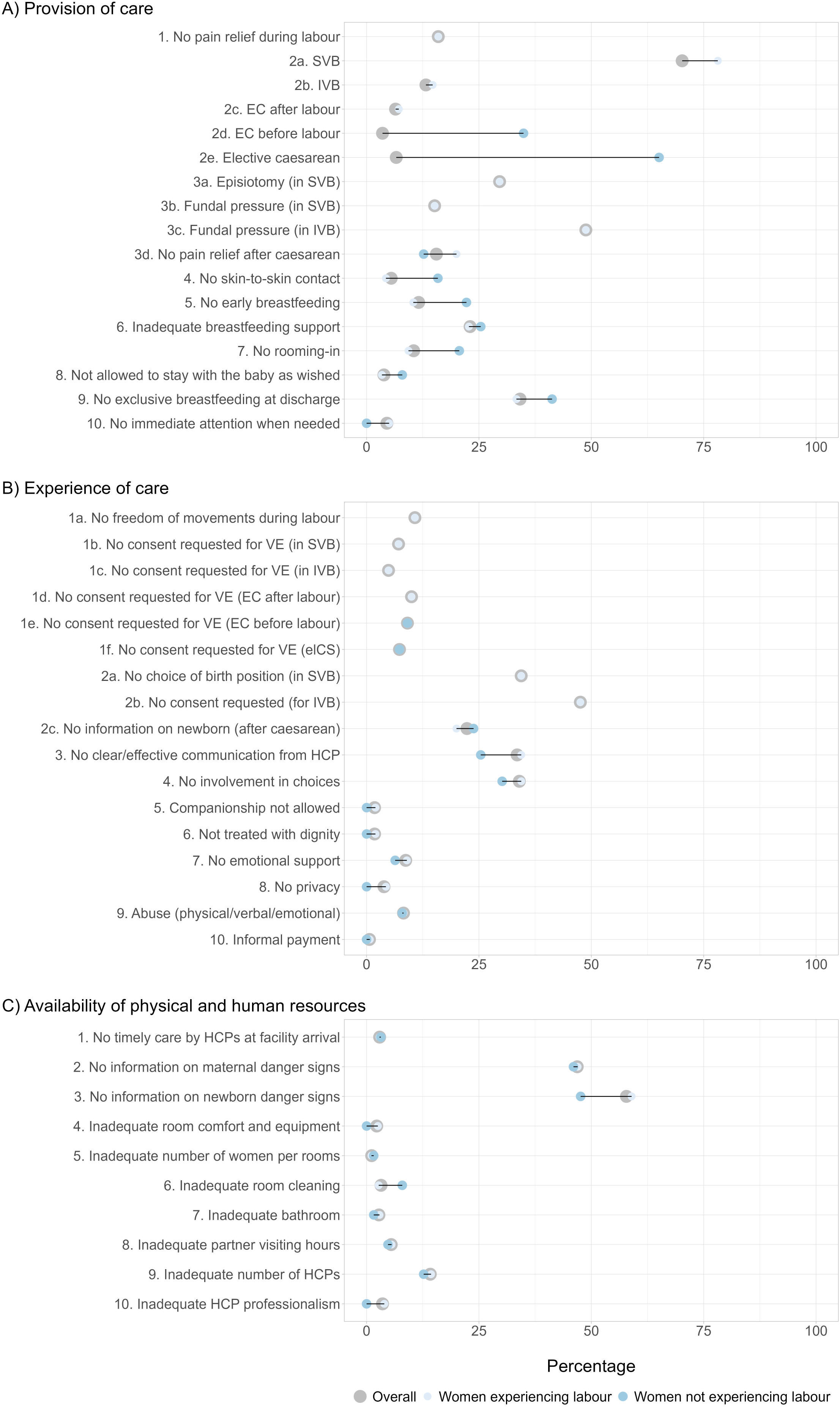
Results for WHO standards-based quality measures, overall and by experience of labour (N=621) Legend: Abbreviations: EC=Emergency Caesarean. elCS=Elective Caesarean. HCP=Health Care Professional. IVB =Instrumental Vaginal Birth. PE=Prelabour Caesarean. SVB=Spontaneous Vaginal Birth. VE=Vaginal Examination.

**Table 2.** Factors influencing emotional childbirth experience assessed by an ordinal logistic regression model (N=480)

|  | Beta coefficient estimate | Standard error | aOR | CI lower bound (2.5%) | CI upper bound (97.5%) | p-value |
| --- | --- | --- | --- | --- | --- | --- |
| **Intercept** |  |  |  |  |  |  |
| Experienced difficulties, with or without effect on mental health | 7.28 | 1.217 |  |  |  | <0.001 |
| **Gave birth in the same country where they were born** |  |  |  |  |  |  |
| No | -1.12 | 0.592 | 0.33 | 0.10 | 1.04 | 0.058 |
| **Region** |  |  |  |  |  |  |
| Brussel region | -0.14 | 0.548 | 0.87 | 0.30 | 2.55 | 0.803 |
| **Age (years)** |  |  |  |  |  |  |
| 18-24 | -0.79 | 0.916 | 0.45 | 0.08 | 2.73 | 0.387 |
| 25-30 | -0.37 | 0.287 | 0.69 | 0.39 | 1.21 | 0.196 |
| 36-39 | -0.21 | 0.401 | 0.81 | 0.37 | 1.78 | 0.597 |
| ≥40 | -1.94 | 1.147 | 0.14 | 0.02 | 1.36 | 0.091 |
| **Education** |  |  |  |  |  |  |
| GCSE (or equivalent) or lower | -0.36 | 0.392 | 0.70 | 0.32 | 1.50 | 0.354 |
| First degree | -0.31 | 0.369 | 0.73 | 0.36 | 1.51 | 0.401 |
| Higher degree (Post-graduate degree/master/doctorate or higher) | -0.04 | 0.306 | 0.96 | 0.53 | 1.76 | 0.906 |
| **Birth mode** |  |  |  |  |  |  |
| Instrumental vaginal birth | 0.86 | 0.391 | 2.36 | 1.10 | 5.09 | 0.028 |
| Emergency caesarean section during labour | 2.70 | 0.541 | 14.94 | 5.17 | 43.16 | <0.001 |
| Emergency caesarean section before going into labour | 2.35 | 0.733 | 10.46 | 2.49 | 44.00 | 0.001 |
| Planned or elective caesarean section before going into labour | 1.00 | 0.643 | 2.71 | 0.77 | 9.58 | 0.121 |
| **Type of healthcare providers who directly assisted birth** |  |  |  |  |  |  |
| Presence of an obstetric doctor in post-graduate training | 0.21 | 0.258 | 1.24 | 0.75 | 2.05 | 0.404 |
| Presence of an obstetric consultant | -0.81 | 0.373 | 0.44 | 0.21 | 0.92 | 0.029 |
| **QMNC index** | -0.03 | 0.004 | 0.97 | 0.96 | 0.98 | <0.001 |
| **Intervention** |  |  |  |  |  |  |
| Episiotomy | 0.06 | 0.306 | 1.07 | 0.58 | 1.94 | 0.836 |
| Fundal pressure | 1.41 | 0.335 | 4.11 | 2.13 | 7.93 | <0.001 |
| **Abuse** |  |  |  |  |  |  |
| Victim of abuse | 1.32 | 0.579 | 3.74 | 1.20 | 11.62 | 0.023 |
| **Parity** |  |  |  |  |  |  |
| ≥2 | -0.07 | 0.279 | 0.94 | 0.54 | 1.62 | 0.815 |

Notes: Abbreviations: CI =confidence interval. aOR=adjusted odds ratio.

Within the domain of provision of care (Figure 2A), 29.6% of women undergoing spontaneous vaginal births reported receiving an episiotomy. Fundal pressure was reported by 15.1% of women with spontaneous vaginal births and by 48.8% of those who had instrumental vaginal births. Among women who underwent a caesarean section, 15.5% reported not receiving pain relief post-operatively. Regarding breastfeeding, 11.6% of all participants did not initiate early breastfeeding, and 34.1% indicated that they did not exclusively breastfeed during hospitalization.

In the domain of experience of care (Figure 2B), about one-third of women reported receiving ineffective or unclear communication from HCPs (33.5%) and an equal proportion reported not being involved in decision-making (34%). Verbal, physical, or emotional abuse was reported by 8.2% of respondents. Additionally, 8.7% reported not receiving emotional support during childbirth, while 3.9% experienced a lack of privacy.

Regarding the availability of resources (Figure 2C), 14.2% of women perceived an inadequate number of HCPs during their stay, and 3.5% reported concerns about the professionalism of healthcare staff. Overall, 46.9% of women did not receive information on maternal danger signs, and 57.8% did not receive information on newborn danger signs. A small proportion (2.9%) reported not receiving timely care upon arrival at the facility, and partner visiting hours were considered inadequate by 5.5% of respondents.

### Emotional childbirth experience and perinatal mental health

Among the 621 women surveyed, more than half (59.1%) reported having a positive and emotionally fulfilling birth experience (Figure 3A), 25.8% indicated they experienced some emotional difficulties that did not significantly affect their overall wellbeing, while 10% reported emotional difficulties that had a negative impact on their wellbeing. Among the 10% of women who reported emotional or psychological difficulties with a negative impact on their wellbeing, the severity of the impact varied: 35.5% described the impact as moderate, another 35.5% as very significant, 27.4% as extreme, and one woman (1.6%) reported a slight impact on her wellbeing (Figure 3D).

**Figure 3.**
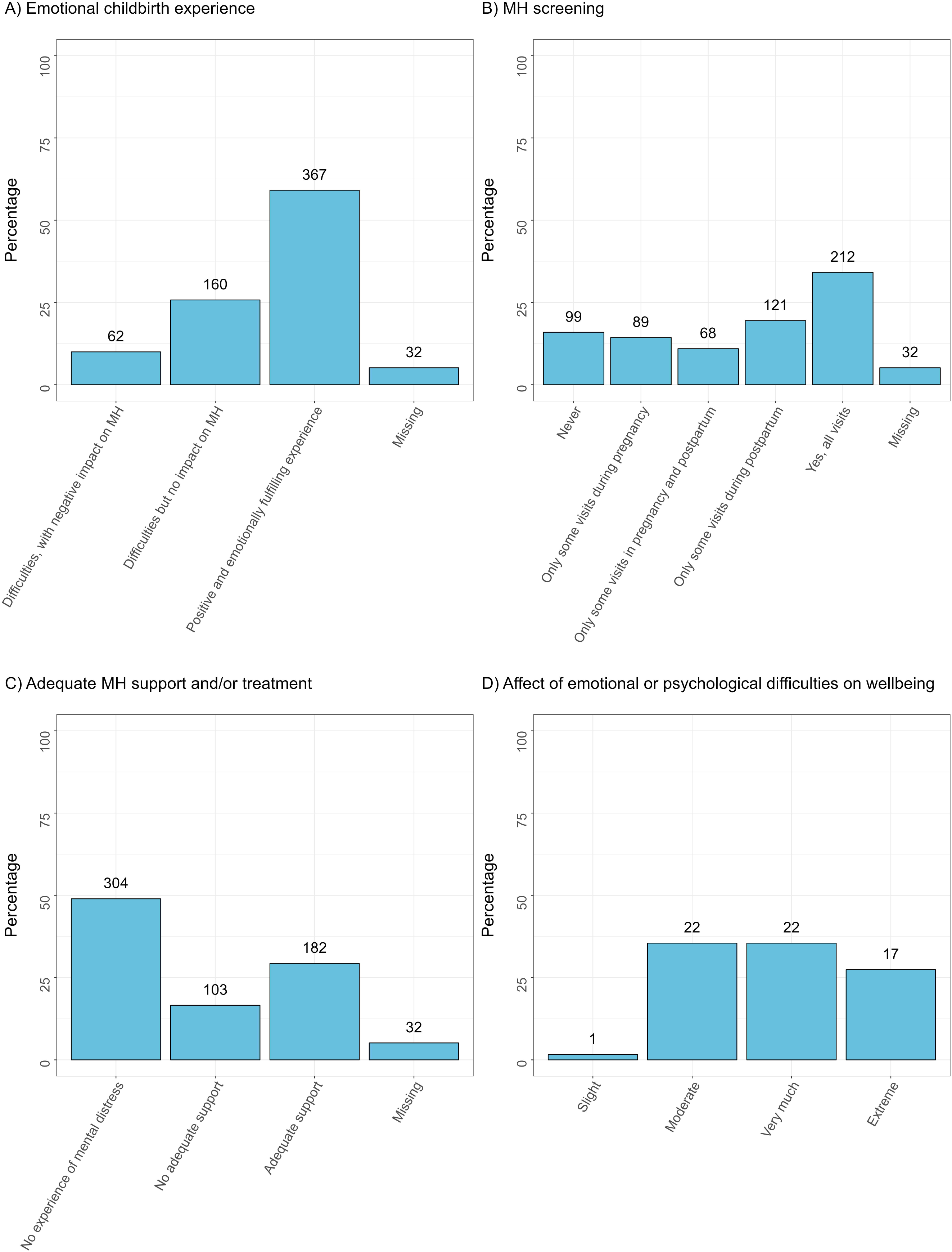
Emotional childbirth experience and mental health screening (N=621) Legend: Numbers above the bars report absolute frequencies. Panel D includes data collected only from the 62 women who reported an emotionally difficult childbirth experience with a negative impact on their mental health, as identified in Panel A Abbreviations: MH = mental health

In terms of mental health screening, one-third of women (34.1%) stated that their mental wellbeing was assessed during all antenatal and postnatal visits (Figure 3B), an additional 14.3% were screened during some antenatal visits, and 10.9% received screening at select times across both the antenatal and postnatal period. Screening occurred only during the postpartum period for 19.5% of participants, while 15.9% reported never being screened for mental wellbeing.

When asked about their emotional state and the support they received, nearly half of respondents (49.0%) indicated they never experienced low mental wellbeing (Figure 3C). Another 29.3% reported mental stress symptoms and received adequate support or treatment, whereas 16.6% reported mental stress symptoms without receiving support to address their mental health needs.

### Quality of care and emotional childbirth experience

Results from the multivariate logistic regression model revealed that mode of birth was significantly associated with emotional difficulties during childbirth. Compared to women who had spontaneous vaginal births, those who had emergency caesarean sections during labour had markedly higher odds of reporting emotional difficulties (aOR: 14.94; 95% CI: 5.17–43.16; p<0.001), as did those who underwent emergency caesarean sections before labour (aOR: 10.46; 95% CI: 2.49–44.00; p=0.001). Instrumental vaginal births were also associated with increased odds of emotional difficulties (aOR: 2.36; 95% CI: 1.10–5.09; p=0.028). Furthermore, we found that women who were subjected to fundal pressure were over four times more likely to report emotional difficulties (aOR: 4.11; 95% CI: 2.13–7.93; p<0.001). Also, those who experienced any form of abuse during childbirth had significantly higher odds of reporting emotional difficulties (aOR: 3.74; 95% CI: 1.20–11.62; p=0.023).

Protective factors for emotional difficulties related to the childbirth experience included the presence of an obstetric consultant during childbirth (aOR: 0.44; 95% CI: 0.21–0.92; p=0.029) and higher overall QMNC index scores. Each point increase in overall QMNC index was associated with reduced odds of emotional difficulties by 3% (aOR: 0.97; 95% CI: 0.96–0.98; p<0.001). Sociodemographic factors such as age, education level, parity, and birthplace of the mother were not significantly associated with reporting emotional difficulties.

## DISCUSSION

This study provides an assessment of the QMNC, the emotional childbirth experience, and perinatal MH of women during the post-COVID-19 period. Furthermore, the relationship between sociodemographic factors, birth characteristics, QMNC and the emotional birth experience was examined.

The reported median QMNC index of 260 reflects overall high-quality standards during childbirth in Belgium. However, despite a high median score, significant shortcomings persist: one-third of women reported ineffective communication and lack of involvement in decisions, while around 9% reported no emotional support from HCPs during childbirth. These findings highlight that the interpersonal dimensions of care remain an area requiring urgent attention. Furthermore, these figures have remained unchanged since the COVID-19 period, indicating that the shortcomings are structural and persistent, rather than a temporary consequence of the pandemic [24]. Another remarkable finding is the high prevalence of fundal pressure, particularly during instrumental births (48.8%), suggesting that this practice remains common despite not being supported by evidence-based guidelines from national organizations, the International Federation of Gynaecology and Obstetrics (FIGO) and WHO [28–30].

The study also examined the emotional birth experience and perinatal MH among women who gave birth in the post COVID-19 period. Alarmingly, over one-third of respondents reported emotional difficulties in the period around childbirth, with 10% noting a negative impact on their wellbeing. Despite national guidelines recommending perinatal MH screening, 16% of women reported never being screened, and among those who experienced low mental wellbeing, a significant proportion (17%) did not receive adequate support [21]. While different initiatives exist to improve perinatal mental healthcare provision (e.g. Born in Belgium Professionals tool, the project “wolk in mijn hoofd” [31, 32]), our findings show a large proportion of women still do not receive the care they need. More research is needed to examine the bottlenecks for adequate MH screening and to determine how to enhance the delivery of MH support in the perinatal period.

Finally, we found that the logistic regression analysis confirms that negative emotional childbirth experiences are associated with certain medical interventions. Emergency caesarean sections (especially during labour) and instrumental vaginal births were strongly associated with emotional distress, consistent with prior research linking emergency interventions to feelings of fear, loss of control, and trauma [33, 34]. Moreover, women who experienced abuse during childbirth were significantly more likely to report emotional difficulties, also a consistent trend in studies on respectful maternity care [35, 36]. Notably, the use of fundal pressure also showed a strong association with negative emotional outcomes, adding to the growing body of evidence pointing to the negative consequences of performing fundal pressure on both medical outcomes and women’s experiences with care [37–39]. Encouragingly, higher QMNC index scores were linked to a lower likelihood of emotional distress, underscoring the importance of high-quality, person-centred care. The presence of an obstetric consultant during birth emerged as a protective factor, suggesting that access to highly skilled health professionals may enhance the overall care experience. However, continuity of care and trust likely play a significant role as well [40, 41]. In Belgium, the obstetric consultant present at birth is usually the same provider who cared for the woman throughout her pregnancy, fostering a strong relationship of trust between the healthcare provider and the woman.

It is important to note that in Belgium maternity care is predominantly obstetric-led, with obstetricians serving as the primary care providers and most births occurring in hospitals (+-99%) [27]. Despite this, research on how to effectively implement person-centred approaches within this medicalised setting remains limited [27]. Such research is urgently needed, especially given that this model is often associated with a high rate of (potentially unnecessary) interventions, affecting the childbirth satisfaction among women [42, 43]. As maternity care becomes increasingly complex, due to rising multimorbidity and greater reliance on technology, ensuring that care remains respectful and centred on the needs of women will continue to be a critical challenge.

### Strengths and Limitations

A major strength of this study is its large and diverse sample, collected through a validated WHO-based tool. The inclusion of MH indicators also offers a novel contribution to maternal health care research in Belgium. However, limitations include the overrepresentation of women speaking Dutch, living in the Flanders Region and women with high education, which may affect the generalizability of findings[44]. Self-selection bias and reliance on self-reported data may also influence results, particularly concerning sensitive issues such as abuse and MH[45].

## Conclusion

This study confirms an overall good level of QMNC in Belgium with gaps in specific quality measures, which are persistent in the post COVID-19 period. Quality of care improvement initiatives should focus on improving adherence to evidence-based guidelines, optimal communication and emotional support, and more structural MH screening. Lastly, more research will be needed to identify and implement strategies that enhance women’s childbirth experiences, especially in relation to medical interventions during childbirth.

## Supporting information

STROBE checklist (cross-sectional study)

Supplementary Box 1. Added mental health questions in the IMAgiNE questionnaire

Supplementary Appendix. Dissemination report

Supplementary Tables 1-3 and model reference categories (additional analyses)

## Data Availability

Data are available upon reasonable request to the corresponding author.

## DECLARATIONS

## Availability of data and materials

Data are available upon reasonable request to the corresponding author.

## Author contributions

ML conceived the IMAgiNE EURO study, with major inputs from MV. AG, JV en VV promoted the survey in Belgium. AG wrote a first draft with input from IM, LGC, and AB for the methods and data analysis. IM and LGC analyzed the data. AG wrote the final draft, with major input from all authors. AG acted as guarantor. All authors approved the final version of the manuscript for submission.

## Acknowledgements

This study received support from the Ministry of Health in Rome, Italy, in partnership with the Institute for Maternal and Child Health IRCCS “Burlo Garofolo” in Trieste. We are grateful to all the women who dedicated their time to complete the survey. We also extend our thanks to our colleagues at the Department of Public Health and Primary Care at Ghent University, the participating maternity hospitals, and everyone who contributed to sharing the survey invitation.

## Disclaimer

The authors are responsible for the views expressed in this article and do not necessarily represent the views, decisions, or policies of the institutions they are affiliated with.

## IMAgiNE EURO Study Group

Austria: Martina König-Bachmann1, Christoph Zenzmaier1, Imola Simon2, Elisabeth D’Costa3, Ursula Kohlendorfer4, Elke Griesmaier4 - 1 Health University of Applied Sciences, Innsbruck, Austria, 2 University of Applied Sciences Burgenland, Austria, 3 Department of Gynecology and Obstetrics, Medical University of Innsbruck, Austria, 4 Department of Pediatrics II, Medical University of Innsbruck, Austria

Belgium: Anna Galle1, Ana Belen Henandez Garcia2, Florence D’Haenens3, Femke Geusens4, Wei Hong Zhang1 - 1 International Centre for Reproductive Health (ICRH) Department of Public Health and Primary Care – Ghent University, 2 Haute Ecole Léonard de Vinci, 3 Hasselt University, Odisee University College of Applied Sciences, 4 KU Leuven University. Bosnia-Herzegovina: Amira Ćerimagić1, Mirha Suceska Nesimovic1 – 1 NGO Baby Steps, Sarajevo, Bosnia-Herzegovina. Chile: Paula Oyarzún Andrades1, Ingrid Vargas Stevenson1, Cristian Carreño Leon, 1 - Unjversidad de Valparaíso. Cyprus: Ourania Kolokotroni1, Eleni Hadjigeorgiou1, Maria Karanikola1, Nicos Middleton1, Ioli Orphanide Eteocleous2 - 1 Cyprus University of Technology School of Health Sciences, 2 Birth Forward NGO. Croatia: Daniela Drandić1,2, Magdalena Kurbanović3, Klaudia Kamenar3 - 1 Roda – Parents in Action, Zagreb, Croatia, 2 International Confederation of Midwives (ICM), 3 Faculty of Health Studies, University of Rijeka, Rijeka, Croatia. Czech Republic: Lenka Laubrova Zirovnicka1, Miloslava Kramná 2- 1 Association For Freestanding Birth Centres and Alongside Midwifery Units (APODAC), 2 Healthy Parenting Association (APERIO). France: Rozée Virginie1, Elise de La Rochebrochard1, Kristina Löfgren2, Sonia Bennis1 - 1 Sexual and Reproductive Health and Rights Research Unit, Institut National d’Études Démographiques (INED), Aubervilliers, France, 2 Baby-friendly Hospital Initiative (IHAB), France. Georgia: Tinatin Manjavidze1, Natia Skhvitaridze1 - University of Georgia. Germany: Céline Miani1, Stephanie Batram-Zantvoort1, Karolina Luegmair2, Anne Kasper3, Mariana Lopes1 - 1 Department of Epidemiology and International Public Health, School of Public Health, Bielefeld University, Bielefeld, Germany, 2 Katholische Stiftungshochschule München University of Applied Sciences 3 University of Applied Sciences and Arts Hildesheim/Holzminden/Göttingen. Greece: Antigoni Sarantaki1, Dimitra Metallinou1, Eirini Orovou2 - 1 Department of Midwifery, School of Health and Care Sciences, University of West Attica, Athens, Greece, 2 Department of Midwifery, University of Western Macedonia, Ptolemaida-Kozani, PC 50200, Greece. Indonesia: Jamilatus Sadiyah1, Gita Nirmala Sari2 - 1 Bumil Pamil Indonesia, 2 Health Polytechnic Ministry of Health of Jakarta III, Indonesia. Israel: Ilana Chertok1,2, Rada Artzi-Medvedik1- 1 Ohio University, School of Nursing, Athens, Ohio, USA, 2 Ruppin Academic Centre, Department of Nursing, Emek Hefer, Israel. Italy: Marzia Lazzerini1, Ilaria Mariani1, Maria Verdecchia1, Margherita Camanni1, Lorenzo Giovanni Cora1, Marianna Zanette1, Mariazzurra Carlino1, Sandra Morano2, Antonella Nespoli3, Simona Fumagalli3, Alessia Melacca4, Nicoletta Di Simone5, Martina Cristodoro5 - 1 Institute for Maternal and Child Health IRCCS Burlo Garofolo, Trieste, Italy, 2 Medical School and Midwifery School, Genoa University, Genoa, Italy, 3 University of Milano Bicocca, Italy, 4 Università degli Studi di Firenze International, Florence, Italy, 5 Humanitas University, Milan, Italy. Latvia: Elizabete Pumpure1, Dace Rezeberga1,2,3, Gunta Lazdane1, Valerija Rakša1, Vaira Avota4, Santa Smilga 1,3, Elīza Meistere 2, 3, Elizabete Rapa 3,2, Arta Berzina 1,3 - 1 Department of Obstetrics and Gynaecology, Rīga Stradiņš University, Rīga, Latvia, 2 Riga East Clinical University Hospital, Rīga, Latvia, 3 Riga Maternity Hospital, Rīga, Latvia, 4 Department of Midwifery, Riga Stradins University, Riga, Latvia. Lithuania: Alina Liepinaitienė 1,2,3, Andželika Šiaučiūnė2, Marija Mizgaitienė4, Simona Jazdauskienė5. Danielė Berulė6 - 1 Department of Environmental Sciences, Faculty of Natural Sciences, Vytautas Magnus University, Kaunas, Lithuania, 2 Lithuanian Midwives Union, Kaunas, Lithuania, 3 SMK College of Applied Sciences, Klaipeda, Lithuania, 4 Kaunas Hospital of the Lithuanian University of Health Sciences, Kaunas, Lithuania, 5 Lithuanian University of Health Sciences, Kaunas, Lithuania, 6 Vilnius University, Vilnius, Lithuania. Luxembourg: Maryse Arendt1 – 1 Beruffsverband vun de Laktatiounsberoderinnen zu Lëtzebuerg asbl (Professional association of the Lactation Consultants in Luxembourg), Luxembourg, Luxembourg, 2 Neonatal intensive care unit, KannerKlinik, Centre Hospitalier de Luxembourg, Luxembourg, Luxembourg. Netherlands: Enrico Lopriore1, Thomas Van den Akker 1,2 - 1 Leiden University Medical Center, Leiden, the Netherlands, 2 Athena Institute, Vrije Universiteit, Amsterdam, Netherlands. Norway: Ingvild Hersoug Nedberg1, Sigrun Kongslien1, Eline Skirnisdottir Vik2 – 1 Department of health and care sciences, UiT The Arctic University of Norway, Norway, 2 Department of health and caring sciences, Western Norway University of Applied Sciences, Norway. Poland: Barbara Baranowska1, Urszula Tataj-Puzyna1, Beata Szlendak1, Paulina Pawlicka2 - 1 Department of Midwifery, Centre of Postgraduate Medical Education, Warsaw, Poland, 2 Division of Intercultural Psychology and Gender Psychology, University of Gdańsk, Gdańsk, Poland. Portugal: Raquel Costa1,2,3, Heloísa Dias4, Tiago Miguel Pinto3, Sofia Marques5,6, Ana Meireles5,6, Joana Oliveira5,6, Mariana Pereira 6, Maria Arminda Nunes 7,8 - 1 EPIUnit - Instituto de Saúde Pública, Universidade do Porto, Rua das Taipas, n° 135, 4050-600 Porto, Portugal, 2 Laboratório para a Investigação Integrativa e Translacional em Saúde Populacional (ITR), Universidade do Porto, Rua das Taipas, n° 135, 4050-600 Porto, Portugal, 3 Lusófona University, HEI-Lab: Digital Human-Environment Interaction Labs, Portugal, 4 Regional Health Administration of the Algarve, (ARS - Algarve, IP), Portugal, 5 Institute of Psychology and Educational Sciences, Lusíada University, Porto, Portugal, 6 CIPD—Psychology for Development Research Centre, Lusíada University, Porto, Portugal, 7 Associação Portuguesa dos Enfermeiros Obstetras, Portugal, 8 Nursing School of Porto, Porto, Portugal Romania: Ana Măiţă1, Ionita-Ciurez Sinziana1 - 1 SAMAS Association, Bucharest, Romania. Serbia: Jelena Radetić1, Jovana Ružičić1 - 1 Centar za mame, Belgrade, Serbia.

Slovenia: Zalka Drglin1, Anja Bohinec1 - 1 National Institute of Public Health, Ljubljana, Slovenia. Spain: Serena Brigidi1, Alejandra Oliden2, Lara Martín Castañeda 1,2 - 1 Institute of Research (VHIR), Vall d’Hebron University Foundation. Maternal and Fetal Medicine Research Group Medical Anthropology Research Center - MARC - Rovira i Virgili University, Tarragona, Spain. Department of Anthropology, Philosophy, and Social Work - Rovira i Virgili University, Tarragona, Spain. President of Observatory of Obstetric Violence in Spain - OVO., 2 Nurse, La casa de Isis Birth Centre - Orba, Alicante, Spain Member of Observatory of Obstetric Violence in Spain – OVO. Sweden: Helen Elden 1,2, Karolina Linden1, Mehreen Zaigham3 - 1 Institute of Health and Care Sciences, Sahlgrenska Academy, University of Gothenburg, Gothenburg, Sweden, 2 Department of Obstetrics and Gynecology, Region Västra Götaland, Sahlgrenska University Hospital, Gothenburg, Sweden, 3 Obstetrics and Gynaecology, Department of Obstetrics and Gynecology, Institution of Clinical Sciences Lund, Lund University, Lund and Skåne University Hospital, Malmö, Sweden. Switzerland: Claire de Labrusse1, Alessia Abderhalden-Zellweger1, Anouck Pfund1 - 1 School of Health Sciences (HESAV), HES-SO University of Applied Sciences and Arts Western Switzerland, Lausanne, Switzerland. Ukraine: Viktoriya Luchka1, Alina Dunayevskaya1 - 1 NGO Safe Birth, Ukraine. United Kingdom: Emile Cote1, Rachel Tribe1, Carlotta Valensin1 - Tribe Lab at King’s College London

