## Supplementary Box 1. Added mental health questions in the IMAgiNE questionnaire for "Quality of maternal and newborn care, perinatal mental health and the emotional birth experience of women: findings of the IMAgiNE EURO study in Belgium"

**ADDITIONAL FILE 2**

Box 1. Added questions on mental health in the IMAgiNE GLOBAL questionnaire

**H8. Did healthcare providers ask you about your mental wellbeing/health, especially in relation to depression and anxiety, during pregnancy and at each appointment post-partum (up to 1 year after childbirth)?**

1) Yes, during all visits both in pregnancy and post-partum

2) Yes, but only in some visits during pregnancy

3) Yes, but only in some visits post-partum

4) No never

**H9. If you felt depressed/hopeless* or anxious/unable to not worry* during pregnancy or after childbirth, do you think that you received adequate help, support and/or treatment (e.g. counselling or another treatment from a competent practitioner*)?**

1) I never felt depressed/hopeless or anxious/unable to not worry

2) I felt depressed/hopeless or anxious/unable to not worry, but I did NOT receive adequate

support

3) I felt depressed/hopeless or anxious/unable to not worry, and I did receive adequate support

**H10. Please select the option that best reflects your experience of childbirth:**

1) I had a positive and emotionally fulfilling birth experience.

2) I experienced emotional or psychological difficulties during childbirth, but overall it did not affect my wellbeing and/or mental health.

3) I experienced emotional or psychological difficulties during childbirth which affected my wellbeing and/or mental health (→10a)

**H10a. How much did these emotional or psychological difficulties affect your wellbeing and/or mental health?**

1) Slightly

2) Moderately

3) Very much

4) Extremely, childbirth was a traumatic experience

**H11. Please suggest 2 actions that could have been taken to improve your mental health/wellbeing**

**during pregnancy and postpartum?**
