## Supplementary Appendix. Dissemination report for "Quality of maternal and newborn care, perinatal mental health and the emotional birth experience of women: findings of the IMAgiNE EURO study in Belgium"

**ADDITIONAL FILE 3**

**Dissemination Report**

Hospitals (13 sites) included a mix of regional and referral hospitals spread over Flanders, mainly through maternity and gynecology wards.

Primary care & midwifery practices (11 sites) spread over Flanders and Brussels included centers of independent midwives, general practitioners, mother & child health centers, and community health centers.

One large daycare center (>100 babies/toddlers) in Flanders was targeted and disseminated the invitation among all parents.

Community & social organizations (6 sites) at national level, such as mutual health funds and professional associations, enabled dissemination on a larger scale.

Online & social media (10 platforms/groups) provided reach via forums, Facebook groups, blogs, and parenting websites.

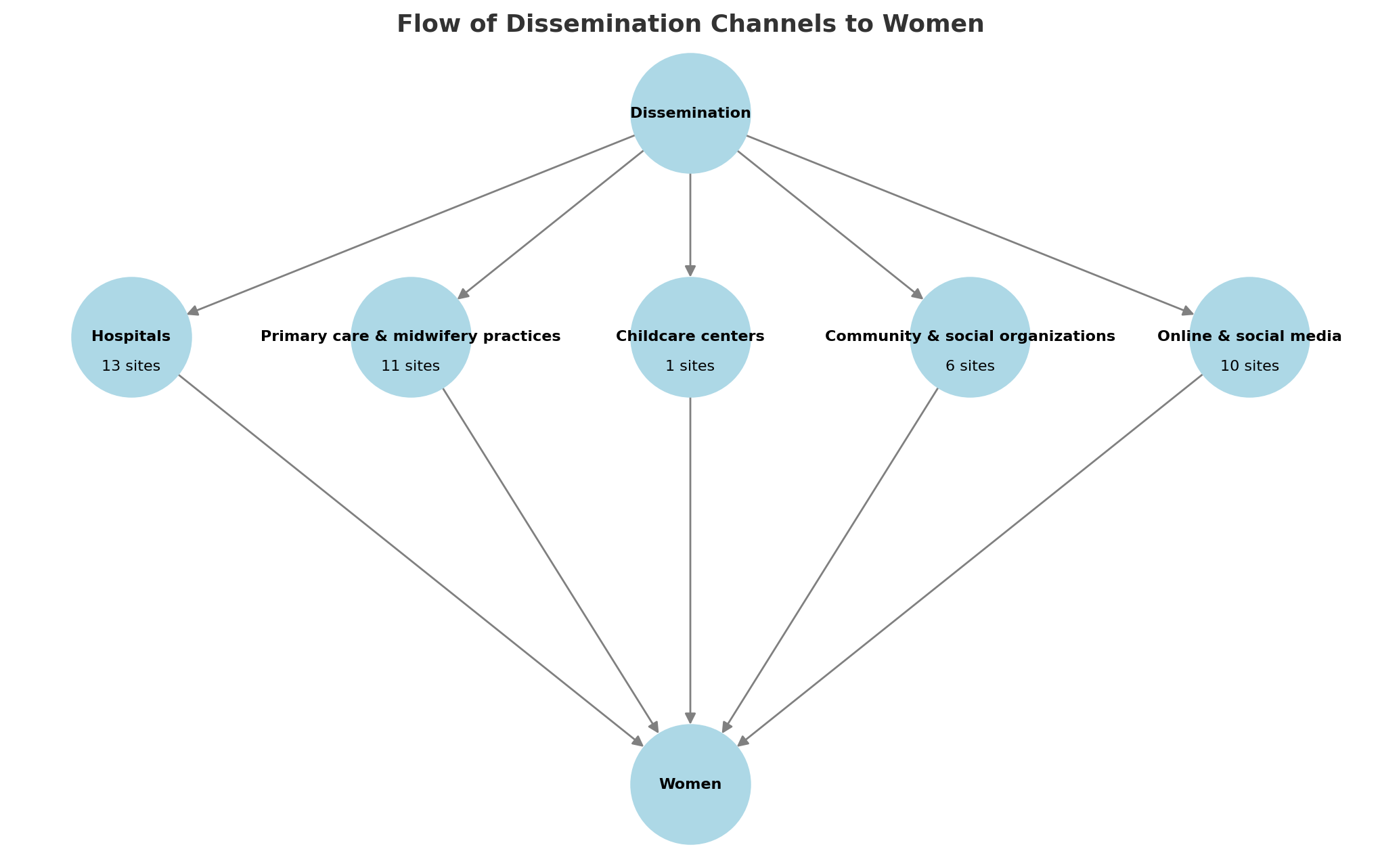
