## Supplementary Tables 1-3 and model reference categories (additional analyses) for "Quality of maternal and newborn care, perinatal mental health and the emotional birth experience of women: findings of the IMAgiNE EURO study in Belgium"

**ADDITIONAL FILE 4**

**Table 1. List of the reference categories for the ordinal logistic regression model**

| **Variable included in the model** | **Reference category** |
| --- | --- |
| **Gave birth in the same country where they were born** | No |
| **Region** | Vlaanderen |
| **Age** | 31-35 years |
| **Education** | Vocational qualification or A level (or equivalent) |
| **Birth mode** | Non-instrumental vaginal birth |
| **Type of healthcare providers who directly assisted birth** | Absence of an obstetric doctor in post graduate training |
|  | Absence of an obstetric consultant |
| **Intervention** | No episiotomy for vaginal birth or caesarean section |
|  | No fundal pressure for vaginal birth or caesarean section |
| **Abuse** | No, never/almost never |
| **Parity** | 1 |

**Table 2. Results of the QMNC Index, overall and by labour experience (N=621)**

|  | **Overall** | | | **Women experiencing labour** | | | **Women not experiencing labour** | | |
| --- | --- | --- | --- | --- | --- | --- | --- | --- | --- |
|  | **N** | **Median** | **IQR** | **N** | **Median** | **IQR** | **N** | **Median** | **IQR** |
| **QMNC Index** | 518 | 260 | [235-275] | 488 | 260 | [235-275] | 30 | 255 | [215-270] |
| **QMNC Index by domain of care** |  |  |  |  |  |  |  |  |  |
| Provision of care | 534 | 90 | [80-95] | 488 | 90 | [80-95] | 46 | 85 | [70-90] |
| Experience of care | 599 | 90 | [75-100] | 558 | 90 | [75-100] | 41 | 90 | [75-100] |
| Availability of physical and human resources | 621 | 80 | [70-90] | 558 | 80 | [70-90] | 63 | 80 | [70-90] |

Notes: The QMNC Index was calculated for women who answered to all the 30 key Quality Measures. The QMNC Index by domain of care was calculated for women who answered to all the 10 key Quality Measures of the domain of care under analysis. The IQR is expressed as [first quartile, third quartile]

Abbreviations: IQR=Interquartile Range. QMNC=Quality of Maternal and Newborn Care.

**Table 3. Results for WHO standards-based quality measures, overall and by experience of labour (N=621)**

|  | **Overall**  (N=621) | | **Women experiencing labour**  (N=558,; %=89.86) | | **Women not experiencing labour**  (N=63; %=10.14) | |
| --- | --- | --- | --- | --- | --- | --- |
| **Provision of care** | **N** | **%** | **N** | **%** | **N** | **%** |
| *Denominator* | *N=558* | | *N=558* | |  |  |
| 1. No pain relief during labour | 89 | 15.95 | 89 | 15.95 | *NA* | *NA* |
| *Denominator* | *N=621* | | *N=558* | |  |  |
| 2a. SVB | 436 | 70.21 | 436 | 78.14 | *NA* | *NA* |
| 2b. IVB | 82 | 13.20 | 82 | 14.70 | *NA* | *NA* |
| 2c. EC after labour | 40 | 6.44 | 40 | 7.17 | *NA* | *NA* |
| *Denominator* | *N=621* | |  |  | *N=63* | |
| 2d. EC before labour | 22 | 3.54 | *NA* | *NA* | 22 | 34.92 |
| 2e. Elective caesarean | 41 | 6.60 | *NA* | *NA* | 41 | 65.08 |
| *Denominator* | *N=436* | | *N=436* | |  |  |
| 3a. Episiotomy (in SVB) | 129 | 29.59 | 129 | 29.59 | *NA* | *NA* |
| 3b. Fundal pressure (in SVB) | 66 | 15.14 | 66 | 15.14 | *NA* | *NA* |
| *Denominator* | *N=82* | | *N=82* | | *NA* | *NA* |
| 3c. Fundal pressure (in IVB) | 40 | 48.78 | 40 | 48.78 | *NA* | *NA* |
| *Denominator* | *N=103* | | *N=40* | | *N=63* | |
| 3d. No pain relief after caesarean | 16 | 15.53 | 8 | 20.00 | 8 | 12.70 |
| *Denominator* | *N=621* | | *N=558* | | *N=63* | |
| 4. No skin-to-skin contact | 34 | 5.48 | 24 | 4.30 | 10 | 15.87 |
| 5. No early breastfeeding | 72 | 11.59 | 58 | 10.39 | 14 | 22.22 |
| 6. Inadequate breastfeeding support | 143 | 23.03 | 127 | 22.76 | 16 | 25.40 |
| 7. No rooming-in | 65 | 10.47 | 52 | 9.32 | 13 | 20.63 |
| 8. Not allowed to stay with the baby as wished | 24 | 3.86 | 19 | 3.41 | 5 | 7.94 |
| 9. No exclusive breastfeeding during hospitalization | 212 | 34.14 | 186 | 33.33 | 26 | 41.27 |
| 10. No immediate attention when needed | 28 | 4.51 | 28 | 5.02 | 0 | 0.00 |
| **Experience of care** |  |  |  |  |  |  |
| *Denominator* | *N=558* | | *N=558* | |  |  |
| 1a. No freedom of movements during labour | 60 | 10.75 | 60 | 10.75 | *NA* | *NA* |
| *Denominator* | *N=436* | | *N=436* | |  | |
| 1b. No consent requested for VE (in SVB) | 31 | 7.11 | 31 | 7.11 | *NA* | *NA* |
| *Denominator* | *N=82* | | *N=82* | |  |  |
| 1c. No consent requested for VE (in IVB) | 4 | 4.88 | 4 | 4.88 | *NA* | *NA* |
| *Denominator* | *N=40* | | *N=40* | |  |  |
| 1d. No consent requested for VE (EC after labour) | 4 | 10.00 | 4 | 10.00 | *NA* | *NA* |
| *Denominator* | *N=22* | |  |  | *N=22* | |
| 1e. No consent requested for VE (EC before labour) | 2 | 9.09 | *NA* | *NA* | 2 | 9.09 |
| *Denominator* | *N=41* | |  |  | *N=41* | |
| 1f. No consent requested for VE (elCS) | 3 | 7.32 | *NA* | *NA* | 3 | 7.32 |
| *Denominator* | *N=436* | | *N=436* | |  |  |
| 2a. No choice of birth position (in SVB) | 150 | 34.40 | 150 | 34.40 | *NA* | *NA* |
| *Denominator* | *N=82* | | *N=82* | |  |  |
| 2b. No consent requested (for IVB) | 39 | 47.56 | 39 | 47.56 | *NA* | *NA* |
| *Denominator* | *N=103* | | *N=40* | | *N=63* | |
| 2c. No information on newborn (after caesarean) | 23 | 22.33 | 8 | 20.00 | 15 | 23.81 |
| *Denominator* | *N=621* | | *N=558* | | *N=63* | |
| 3. No clear/effective communication from HCPs | 208 | 33.49 | 192 | 34.41 | 16 | 25.40 |
| 4. No involvement in choices | 211 | 33.98 | 192 | 34.41 | 19 | 30.16 |
| 5. Companionship not allowed | 11 | 1.77 | 11 | 1.97 | 0 | 0.00 |
| 6. Not treated with dignity | 11 | 1.77 | 11 | 1.97 | 0 | 0.00 |
| 7. No emotional support | 54 | 8.70 | 50 | 8.96 | 4 | 6.35 |
| 8. No privacy | 24 | 3.86 | 24 | 4.30 | 0 | 0.00 |
| 9. Abuse (physical/verbal/emotional) | 51 | 8.21 | 46 | 8.24 | 5 | 7.94 |
| 10. Informal payment | 4 | 0.64 | 4 | 0.72 | 0 | 0.00 |
| **Availability of physical and human resources** |  |  |  |  |  |  |
| *Denominator* | *N=621* | | *N=558* | | *N=63* | |
| 1. No timely care by HCPs at facility arrival | 18 | 2.90 | 16 | 2.87 | 2 | 3.17 |
| 2. No information on maternal danger signs | 291 | 46.86 | 262 | 46.95 | 29 | 46.03 |
| 3. No information on newborn danger signs | 359 | 57.81 | 329 | 58.96 | 30 | 47.62 |
| 4. Inadequate room comfort and equipment | 14 | 2.25 | 14 | 2.51 | 0 | 0.00 |
| 5. Inadequate number of women per rooms | 7 | 1.13 | 6 | 1.08 | 1 | 1.59 |
| 6. Inadequate room cleaning | 20 | 3.22 | 15 | 2.69 | 5 | 7.94 |
| 7. Inadequate bathroom | 17 | 2.74 | 16 | 2.87 | 1 | 1.59 |
| 8. Inadequate partner visiting hours | 34 | 5.48 | 31 | 5.56 | 3 | 4.76 |
| 9. Inadequate number of HCPs | 88 | 14.17 | 80 | 14.34 | 8 | 12.70 |
| 10. Inadequate HCPs professionalism | 22 | 3.54 | 22 | 3.94 | 0 | 0.00 |

Abbreviations: EC=Emergency Caesarean. elCS=Elective Caesarean. HCP=Health Care Professional. ICB=Instrumental Vaginal Birth. NA=Not Applicable. PE=Prelabour Caesarean. SVB=Spontaneous Vaginal Birth. VE=Vaginal Examination.
